# Social Determinants of Health and Fatal Crashes Involving US Geriatric and Non-Geriatric Road Users

**DOI:** 10.1101/2023.06.23.23291843

**Authors:** Oluwaseun Adeyemi, Charles DiMaggio, Corita Grudzen, Sanjit Konda, Erin Rogers, Keith Goldfeld, Saul Blecker, Joshua Chodosh

## Abstract

**Introduction:** Social determinants of health (SDoH), defined as nonmedical factors that impact health outcomes, have been associated with fatal crash occurrences. Road users who live in communities with negative SDoH may be at increased risk of crash-related mortality, and the risks may be further heightened among geriatric road users and in rural areas. We evaluated the relationship between the county-level measure of SDoH and county-level fatal crash counts among geriatric and non-geriatric road users living in rural, suburban, and urban areas.

**Methods:** For this ecological study, we pooled data from Fatality Analysis Reporting System (2018 to 2020) and the U.S. Census Bureau (2019 data) and limited our analyses to the 3,108 contiguous US counties. The outcome measures were county-level fatal crash counts involving (1) geriatric (65 years and older) road users (2) non-geriatric road users, and (2) the general population. The predictor variable was the Multidimensional Deprivation Index (MDI), a score that measures the five domains of SDoH - economic quality, healthcare access, education, community, and neighborhood quality. We defined the MDI as a three-level categorical variable: at or below the national average, within two-fold of the national average, and higher than two-fold of the national average. We controlled for county-level demographics and crash characteristics. We performed a Bayesian spatial Poisson regression analysis using Integrated Nested Laplace Approximations and reported the crash fatality rate ratios (plus 95% Credible Intervals (CrI)).

**Results:** The median (Q1, Q3) standardized mortality rate ratios among geriatric and non-geriatric road users were 1.3 (0.6, 2.5) and 1.6 (0.9, 2.7), respectively. A total of 283 (9.1%) and 806 (15.9%) counties were classified as very highly deprived and highly deprived, respectively. Clusters of counties with high deprivation rates were identified in the Southern states. Counties classified as very highly deprived and highly deprived had 40% (95% CrI: 1.24 – 1.57) and 25% (95% CrI: 1.17 – 1.34) increased geriatric fatality crash rate ratios and this pattern of association persisted in suburban and urban areas. Also, counties classified as very highly deprived and highly deprived had 42% (95% CI: 1.27 – 1.58) and 32% (95% CI: 1.23 – 1.38) increased fatality crash rate ratios among all road users and this pattern persisted in suburban and urban areas. Counties with more than four-fold increased fatality rate ratios were located commonly in Texas, Oklahoma, Nevada, and Utah.

**Conclusion:** Despite older adults being less frequent road users, county-level deprivation measures of the SDoH are equally associated with geriatric and non-geriatric crash-related fatal rate ratios. Policies that improve county-level SDoH may reduce the county-level fatal rate ratios equally among geriatric and non-geriatric road users.

## Introduction

Social determinants of health are conditions located where people live that affect people’s quality of life and health outcomes.^1-3^ These conditions are commonly grouped into five interrelated domains: (1) economic stability, (2) education access and quality, (3) healthcare access and quality, (4) neighborhood and built environment, and (5) community and social context.^3^ Each of these wide-ranging conditions has been associated with individual and community-level health disparity and inequities and may serve as predictors of increased susceptibility to acute and chronic disease morbidity and mortality. Poverty, an exemplar of economic stability, predicts substance use, injury, and adverse events from chronic diseases.^4^ Similarly, rurality, an exemplar of neighborhood and built environment is associated with increased hospital closure,^5-7^ longer emergency medical service response and hospital transit time,^8-11^, and increased deaths without resuscitation.^10^

Understanding the relationship between SDoH and fatal crash counts will further highlight the importance of improving measures of SDoH. The literature on geriatric fatal crash injuries is sparse and very few studies assess the relationship between SDoH and crash injuries.^12-15^ Yet, SDoH may provide insight into how the conditions surrounding the place we live influence the occurrence of crash injuries. This study, therefore, fills the gap in the SDoH and crash injury literature using three sets of objectives. First, we identified counties with high clusters of negative SDoH and fatal crash rates in the geriatric and non-geriatric populations. We hypothesize that there will be a heterogenous distribution of SDoH and fatal crash rates across the US. Secondly, we assessed the relationship between SDoH and fatal crash counts across the geriatric and non-geriatric populations. We hypothesized that county-level measure of SDoH will be associated with county-level fatal crash counts and the pattern of association will not differ among the geriatric and non-geriatric populations. Lastly, we hypothesize that the effect size of the association between SDoH and geriatric and non-geriatric fatal crash counts will be greater in rural areas compared to urban areas.

## Methods

### Study Design and Population

For this ecological study, we employed a cross-sectional design to assess the association between county-level measures of fatal crash counts and SDoH. Our unit of analysis was at the county-level

### Outcome Variable

The outcome variable is county-level fatal crash counts. We extracted data on fatal crash counts from the Fatality Analysis Reporting System (FARS), one of the national crash databases of the National Highway Traffic Safety Administration.^16, 17^ The FARS dataset is a census of crash events during which one or more persons died either at the scene or within 30 days of the index crash event.^16^ The FARS data consists of multiple linkable files that provide information on the accident, person, vehicle, and risk factor-related information. Data in the FARS database is provided either at the individual or crash event levels.

For this study, we defined three sets of county-level fatal crash deaths - across all road users, geriatric (aged 65 years and older), and non-geriatric road users. We defined these three sets of county-level fatal crash counts in three steps. First, from the person file of the FARS data, we created three datasets, restricting each by age (dataset 1: all ages, dataset 2: 65 years and older, dataset 3: less than 65 years).

Next, we restricted each dataset to persons whose injury status was recorded as dead. Lastly, we aggregated the death counts in each dataset by county using the five-digit Federal Information Processing System (FIPS) code. We limited the counties to the 3,108 contiguous US counties, excluding Alaska and the Virgin Islands.

### Predictor Variable

The predictor variable is the multidimensional deprivation index (MDI). The MDI is a measure of SDoH developed by the US Census Bureau.^18^ It consists of six dimensions: standard of living, health, education, economic security, housing quality, and neighborhood quality (Report, A.C.S., Multidimensional Deprivation in the United States: 2017. 2019.).^19^ Each of the six dimensions is measured at the individual level and has specific defining criteria.^19^ An individual is considered deprived if at least two criteria are met. The ACS published the national and county-level deprivation index,^18^ constructed from individual-level measures using the Alkire Foster method.^20^ Additionally, the ACS defined MDI was defined as a five-level categorical variable defined as county deprivation - very low (i.e. less than half of the national rate), low (ranged from just above half of the national rate and below the lower limit of the 90% confidence interval), at national level (within 90% confidence interval of the national rate), high (above the upper limit of the 90% confidence interval and just below twice the national rate), and very high (at least two times the national rate).^18^ For this study, we defined MDI as a continuous measure and a three-level categorical variable - very low, low, at the national average, or higher.

### Control Variable

We selected county-level sociodemographic and crash characteristics based on prior literature. The county-level sociodemographic and health characteristics included proportions of males, Blacks, Hispanics, the poor, those with bachelor’s degrees, and excessive alcohol intake and emergency department utilization rate. These variables, excluding excessive alcohol intake and emergency department utilization rate, were extracted from American Community Survey.^21^ Excessive alcohol intake, defined as the proportion of adults with self-reported binge or heavy drinking, was extracted from the County Health Rankings and Roadmaps.^22, 23^ The emergency department utilization per 1000 Medicare beneficiaries was extracted from the Agency for Healthcare Research and Quality.^24^ The county-level crash characteristics include the proportions of fatal crash events that occurred at night, during the rush hour period, and the proportion of crash victims screened for alcohol and drugs. Night driving was defined as crash events occurring between 12 midnight and 6 am. Rush hour driving was defined as crash events occurring between 6 and 9 am, and 3 and 7 pm.^25^ Persons with positive alcohol use were defined as those with a positive blood alcohol level of 0.08% or higher or whose alcohol use status was either self-reported or officer-reported. Persons with positive drug use were defined as those with positive drug tests or whose drug status was either self-reported or officer-reported. All crash characteristics were extracted from FARS.

### Stratification

We stratified the counties by rurality/urbanicity using the 2010 Rural-Urban Commuting Area (RUCA) codes (ref).^26^ The RUCA code is a 10-level classification based on density, urbanization, and daily commuting. We defined rurality/urbanicity in three levels: metropolitan-urban (RUCA codes 1-3), micropolitan-urban (RUCA codes 4-7), and rural and small towns (RUCA codes 8-10).

### Spatial Weight Matrix

Using the Euclidean distance across the 3,108 contiguous US counties, we created a spatial weight matrix to map the spatial relationship across the counties. We used the inverse-weighted interpolation, which defines weight as a decreasing function of distance, to generate the spatial weight and account for the presence of spatial autocorrelation in crash occurrences.^27^

## Data Analysis

We reported the proportions of county-level fatal crash injury metrics (counts, case fatality rates (CFR), and standardized mortality ratio (SMR)) across all road users and among geriatric and non-geriatric road users in metropolitan-urban, micropolitan-urban, and rural areas. We reported the frequency distributions, mean (and standard deviation (SD)), and median (and first and third quartiles (Q1, Q3)) for all the predictor and control variables as appropriate. We visualized the distribution of the county-level crude fatality rates and MDI values on choropleths. We assessed the construct validity of the MDI across the crash, health, and population characteristics using correlation analysis. MDI was adjudged to correlate with variables across the domains of SDoH if the correlation coefficient was ≥0.3. For the regression models, variables with correlation coefficients of ≥0.3 were deemed to exhibit substantial correlation with MDI and were removed from the models.

Since our outcome variable is a count measure, we performed model testing to compare the Poisson, negative binomial regression, and zero-inflated Poisson and negative binomial regression models, using a spatial Bayesian paradigm with non-informative priors. The offset variable was the county-level expected count, defined as county population multiplied by the ratio of the sum of fatal counts and the sum of the population across the 3108 counties. Consistent with our definition of the outcome variable, we generated three different county population sizes (all ages, geriatric, and non-geriatric), and computed county-level expected counts for all road users, and geriatric and non-geriatric road users. We reported the Akaike Information Criteria and Bayesian Information Criteria.

After performing model fitting and selecting the Poisson model, we performed a spatial Bayesian Poisson regression analysis. For the actual fatal crash counts *Y(s*_*i*_),we assumed a Poisson distribution in the data. The mortality rate ratio {λ(*S*_*i*_),*i* = 1,…,*n*} over the n = 3, 108 counties were modeled by county-level K independent variables *X*_*k*_*(S*_*i*_*),k = 1, …, K*,, a set of regression coefficients *β*_*k*_,*K = 0,1, …, K*,, spatial random effects terms {*V(S*_*i*_*),i = 1, …, n*}, and a error components {*ε(S*_*i*_*), i = 1, …, n*} that follows independent and identical zero-mean Gaussian distributions with variance parameter σ^2^. The regression equation takes the form:

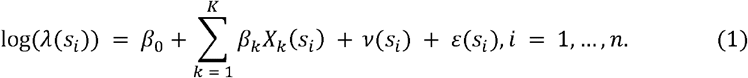

Thereafter, we reported the unadjusted and adjusted mortality rate ratios (incidence rate ratios) and reported the 95% credible interval. For each model, we computed the mortality rate ratios for all road users as well as geriatric and non-geriatric road users. We computed the predicted rate ratios for each county and visualized the distribution on choropleths. Additionally, we performed stratified analysis and for each of the three outcome measures, we generated mortality rate ratios by rurality/urbanicity.

## Results

Across the 3,108 counties, the median (Q1, Q3) MDI rate was 13.0 (9.4, 18.1) (Table 1). The proportion of counties classified as high and very highly deprived were 9% and 26%, respectively. The mean (SD) crash case fatality rate across all age groups was 44.7% (15.1), with the rates lower among the geriatric population (9.1% (8.8)) compared to the non-geriatric population (35.6% (1.0)). The median (Q1, Q3) county SMR across all age groups was 1.6 (1.0, 2.7), with the ratio marginally lower among the geriatric 1.3 (0.6, 2.5) population compared to the non-geriatric population 1.6 (0.9, 2.7). The mean county proportion of males was 50% (2.2) while the median county proportions of non-Hispanic Blacks and Hispanics were 2.6% and 4.5%, respectively. The mean county proportion of the poor classified as poor was 14.5% while those with bachelor’s degree was 19.6%. Also, the mean county proportions of alcohol intake and ED utilization rates were 0.2% and 5.6%, respectively. Approximately a quarter of crashes occurred at night and 43% occur during the rush hour period. Furthermore, mean alcohol and drug screening rates across the counties were 43% and 39%, respectively. Counties with very high deprivation index were found in South Dakota, Arizona, New Mexico, and in some southern region states - Texas, Louisiana, Mississippi, Alabama, Georgia, South Carolina, and Kentucky (Figure 1a). County clusters of high deprivation index were similarly identified in South Dakota, Arizona, New Mexico, and in all southern US states (Figure 1b). Counties with standardized mortality rates of 10 or higher were identified in Montana, North and South Dakota, Wyoming, Colorado, New Mexico, Texas, Nevada, California, and Oregon (Figure 2a). This pattern of high standardized mortality rate was consistent among geriatric and non-geriatric road users (Figure 2b and 2c).

**Table 1:** County-level frequency distribution and summary statistics of multidimensional deprivation index, crash, and sociodemographic characteristics by rurality/urbanicity (N=3,108)

| Variable | All<br>(N=3,108) | Rural + Small Towns<br>(n=627 (20.2%)) | Micropolitan-Urban<br>(n=1,321 (42.5%)) | Metropolitan-Urban<br>(n=1,160 (37.3%)) | p-value |
| --- | --- | --- | --- | --- | --- |
| Multidimensional Deprivation Index |  |  |  |  |  |
| MDI Rate (Median (Q1, Q3)) | 13.0 (9.4, 18.1) | 13.4 (9.7, 19.0) | 14.9 (10.8, 19.3) | 11.4 (8.1, 15.5) | <0.001<sup>##</sup> |
| Deprivation Index (n (%)) |  |  |  |  |  |
| Very highly deprived | 283 (9.1) | 125 (11.4) | 109 (11.8) | 49 (4.5) | <0.001<sup>###</sup> |
| Highly deprived | 806 (25.9) | 290 (26.5) | 302 (32.6) | 214 (19.7) |  |
| Average to Low Deprived | 2,019 (65.0) | 681 (62.1) | 516 (55.7) | 822 (75.8) |  |
| Fatal Crash Measures |  |  |  |  |  |
| Fatal Counts (Geriatric) (Median (Q1, Q3)) | 3.0 (1.0, 6.0) | 1.0 (0.0, 2.0) | 3.0 (1.0, 5.0) | 6.0 (3.0, 11.0) | <0.001<sup>##</sup> |
| Fatal Counts (Non-Geriatric) (Median (Q1, Q3)) | 11.0 (5.0, 23.0) | 5.0 (2.0, 8.0) | 13.0 (7.0, 20.0) | 25.0 (11.0, 50.0) | <0.001<sup>##</sup> |
| Fatal Counts (All Ages) (Median (Q1, Q3)) | 14.0 (6.0, 28.0) | 6.0 (3.0, 11.0) | 16.0 (9.0, 25.0) | 31.0 (15.0, 62.0) | <0.001<sup>##</sup> |
| Case Fatality Rate (Geriatric) (Mean (SD)) | 9.1 (8.8) | 10.3 (12.3) | 8.8 (6.8) | 8.2 (5.7) | <0.001<sup>#</sup> |
| Case Fatality Rate (Non-Geriatric) (Mean (SD)) | 35.6 (1.0) | 38.8 (19.6) | 35.1 (11.5) | 32.9 (10.5) | <0.001<sup>#</sup> |
| Case Fatality Rate (All Ages) (Mean (SD)) | 44.7 (15.1) | 49.1 (19.0) | 43.9 (12.6) | 41.1 (11.2) | <0.001<sup>#</sup> |
| SMR (Geriatric) (Median (Q1, Q3)) | 1.3 (0.6, 2.5) | 1.7 (0.0, 3.5) | 1.5 (0.9, 2.5) | 1.1 (0.6, 1.7) | <0.001<sup>##</sup> |
| SMR (Non-Geriatric) (Median (Q1, Q3)) | 1.6 (0.9, 2.7) | 2.3 (1.3, 3.7) | 1.8 (1.2, 2.6) | 1.1 (0.7, 1.7) | <0.001<sup>##</sup> |
| SMR (All Ages) (Median (Q1, Q3)) | 1.6 (1.0, 2.7) | 2.3 (1.4, 3.6) | 1.8 (1.2, 2.6) | 1.1 (0.7, 1.7) | <0.001<sup>##</sup> |
| Sociodemographic Characteristics |  |  |  |  |  |
| Male proportion (Mean (SD)) | 50.1 (2.2) | 50.6 (2.6) | 50.1 (2.2) | 49.5 (1.6) | <0.001<sup>#</sup> |
| Black proportion (Median (Q1, Q3)) | 2.6 (0.9, 11.1) | 1.0 (0.6, 3.5) | 2.8 (1.1, 11.4) | 6.4 (1.9, 15.8) | <0.001<sup>##</sup> |
| Hispanic proportion (Median (Q1, Q3)) | 4.5 (2.5, 10.2) | 3.5 (2.1, 7.3) | 4.4 (2.5, 10.6) | 5.8 (3.1, 12.1) | <0.001<sup>##</sup> |
| Poverty proportion (Mean (SD)) | 14.5 (5.8) | 15.6 (6.3) | 15.8 (5.6) | 12.2 (4.7) | <0.001<sup>#</sup> |
| Bachelor's degree proportion (Median (Q1, Q3)) | 19.6 (15.3, 26.0) | 18.1 (14.3, 22.2) | 17.5 (14.3, 22.1) | 25.2 (18.7, 33.5) | <0.001<sup>##</sup> |
| Excessive alcohol intake proportion (Mean (SD)) | 0.2 (0.1) | 0.2 (0.1) | 0.2 (0.1) | 0.2 (0.1) | <0.001<sup>#</sup> |
| ED utilization rate (Mean (SD)) | 5.6 (1.1) | 5.5 (1.3) | 6.0 (1.0) | 5.4 (0.9) | <0.001<sup>#</sup> |
| Crash Event Characteristics |  |  |  |  |  |
| Night fatal crash proportion (Mean (SD)) | 24.8 (17.3) | 21.5 (21.7) | 24.5 (14.4) | 28.4 (13.6) | <0.001<sup>#</sup> |
| Rush hour fatal event proportion (Mean (SD)) | 43.3 (20.9) | 43.0 (28.2) | 44.5 (16.8) | 42.7 (14.2) | 0.112 |
| Alcohol screening proportion (Mean (SD)) | 43.3 (20.2) | 46.1 (25.2) | 42.7 (17.5) | 40.9 (16.2) | <0.001<sup>#</sup> |
| Drug screening proportion (Mean (SD)) | 38.7 (22.9) | 38.7 (26.9) | 38.8 (21.3) | 38.8 (19.7) | 0.995 |

SMR: Standardized Mortality Rate.; CFR: Case Fatality Rate; ED: Emergency Department; #: One-way ANOVA performed; ##: Kruskal-Wallis test performed; ###: Chi-square test performed

**Table 2:**
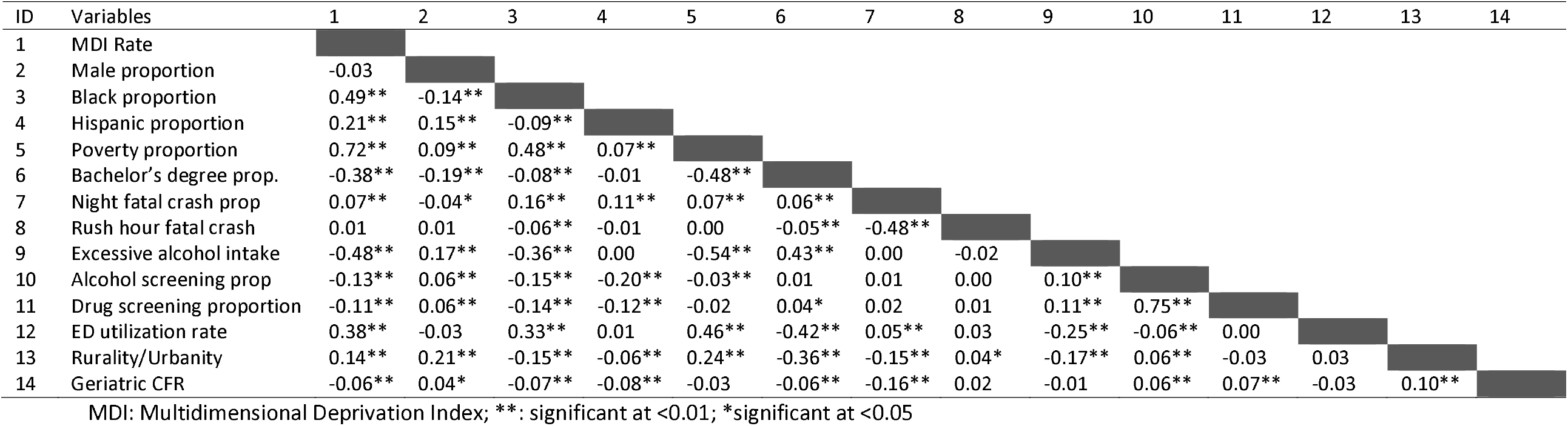
Pairwise correlation coefficient assessing the relationship between county-level multidimensional deprivation index, sociodemographic characteristics and fatal crash events

**Figure 1:**
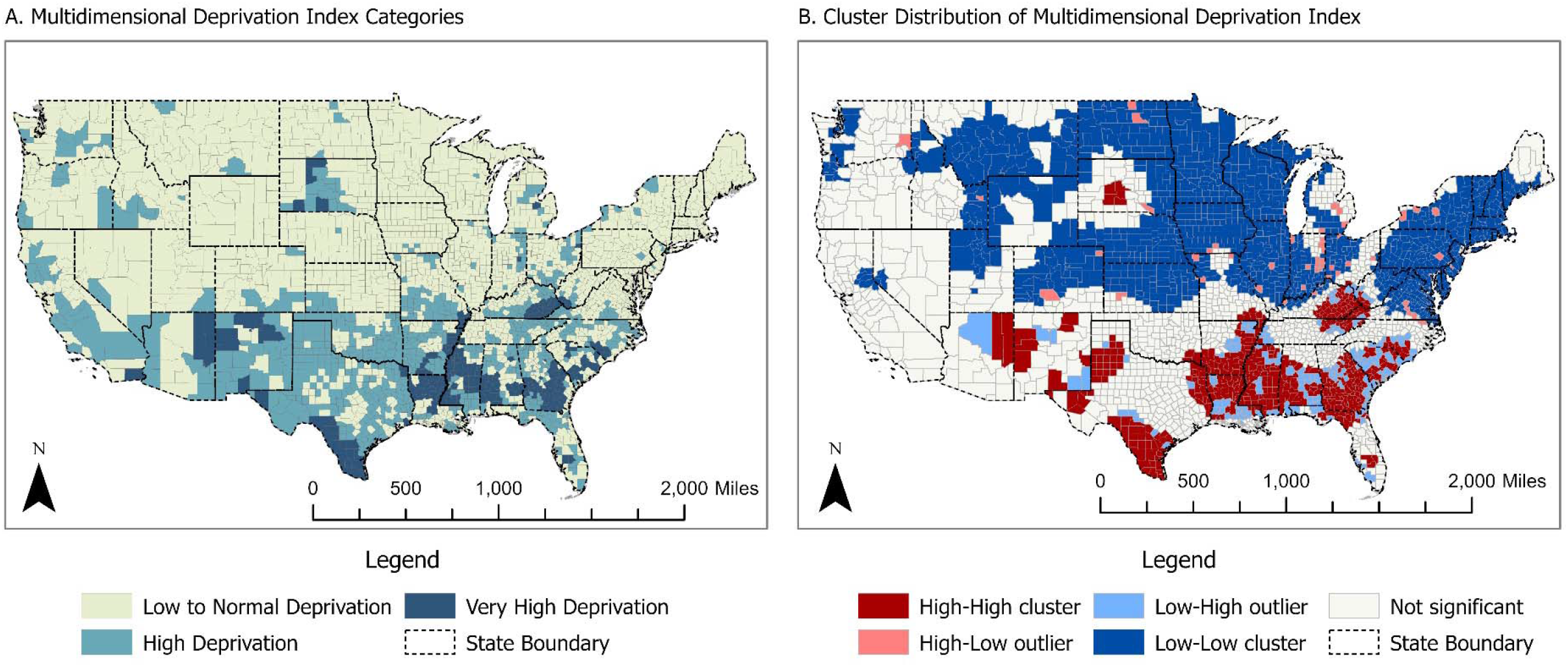
A. U.S. County distribution of the Multidimensional Deprivation Index B. Cluster identification of multidimensional deprivation index across U.S. counties

**Figure 2:**
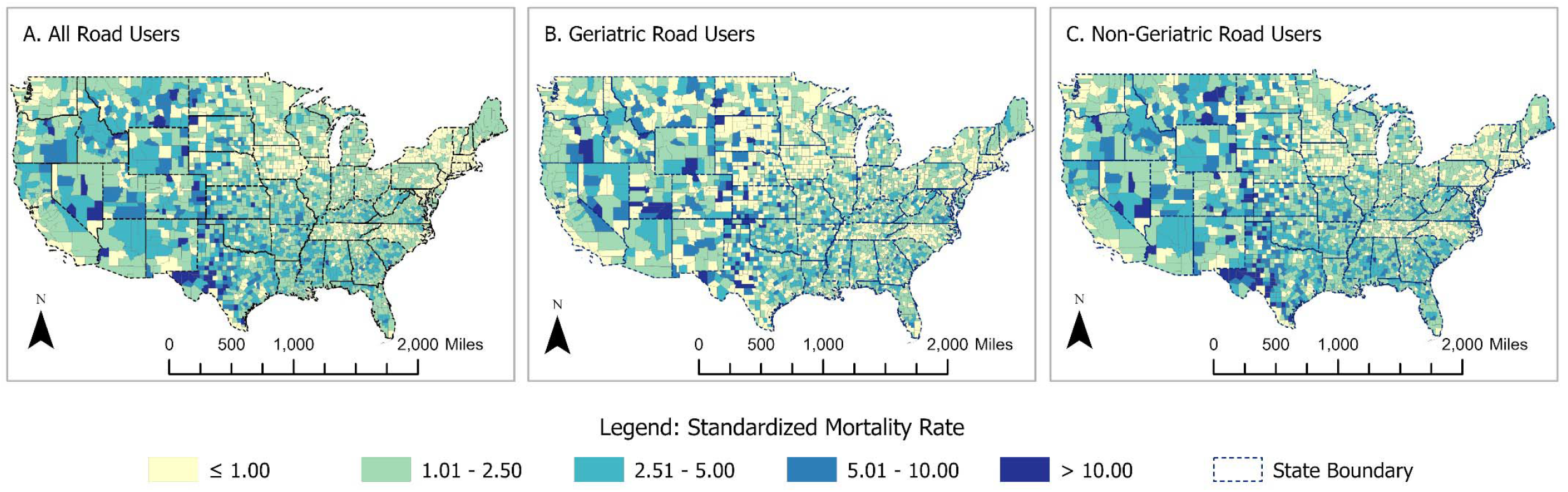
County-level distribution of the motor vehicle crash standardized mortality rates among (A) road users of all ages (B) geriatric road users, and (C) non-geriatric road users.

We evaluated the construct validity of the MDI across the five constructs of the SDoH. In the economic stability domain, MDI exhibited a statistically significant strong positive correlation with the county proportions of those classified as poor (r=0.72, p<0.01). In the education access and quality domain, MDI exhibited statistically significant moderate negative correlations with county proportions of those with bachelor’s degrees (r= -0.38, p<0.01). In the healthcare access and quality domain, MDI exhibited moderate positive correlations with county-level ED utilization rate (r= 0.38, p<0.01). In the neighborhood and built environment, MDI exhibited a statistically significant mild positive correlation with rurality (r=0.14; p<0.01). In the community and social context domain, MDI exhibited a statistically significant moderate positive correlation with county proportions of Blacks (r=0.49, p<0.01) and a moderate negative correlation with county proportions of excessive alcohol intake (r=-0.48, p<0.01).

The result of the model testing showed that the spatial Poisson regression was the most parsimonious model compared to the negative binomial, zero-inflated negative binomial, and zero-inflated Poisson models (Table 3). Across the entire population and separately among the geriatric and non-geriatric populations, the spatial Poisson model had the lowest AIC and DIC.

**Table 3:** Summary of Model Testing

| Equation | $Y = \beta_0 + \beta_1 VHD + \beta_2 MD2 + \beta_3 Covariates + Exp$ (Expected Counts)<br>+ (1\| *County ID*) + (1\| *Spatial Weight Matrix*) | | |
| --- | --- | --- | --- |
| Categories | Models | DIC | WAIC |
| Geriatric Road Users | Poisson | 12773.08 | 12797.04 |
|  | Negative Binomial | 13078.22 | 13097.60 |
|  | Zero-inflated Poisson | 12799.53 | 12837.25 |
|  | Zero-inflated Negative Binomial | 13099.47 | 13141.36 |
| Non-Geriatric Road Users | Poisson | 18272.93 | 18148.86 |
|  | Negative Binomial | 18862.41 | 18670.81 |
|  | Zero-inflated Poisson | 18306.93 | 18226.43 |
|  | Zero-inflated Negative Binomial | 18791.44 | 18766.62 |
| All Road Users | Poisson | 19123.49 | 18972.55 |
|  | Negative Binomial | 19596.30 | 19490.90 |
|  | Zero-inflated Poisson | 19126.44 | 18972.53 |
|  | Zero-inflated Negative Binomial | 19418.82 | 19319.98 |

Y represents Y1 (fatal counts of geriatric road users) or Y2 (fatal counts of non-geriatric road users) or Y3 (fatal counts of all road users); VHD: Dummy of Very Highly Deprived vs. Average to Low Deprived; HD: Dummy of Highly Deprived vs. Average to Low Deprived; Exp: Exposure; DIC: Deviance Information Criteria; WAIC: Watanabe-Akaike Information Criteria; Expected counts computed as: [Sum (Fatal counts)/ Sum (County Population)] \* County Population

Across all age groups, a unit increase in MDI was associated with a 1% increase in crash fatality rates (Table 4). Also, those living in very highly deprived and highly deprived counties had 33% (95% CrI: 1.20 - 1.48) and 25% (95% CrI: 1.17 - 1.32) increased crash mortality rate ratio compared to those living in counties with normal MDI rate. This pattern of association was consistent among geriatric and non-geriatric road users. Several states in the West and South of the US had four-fold crash mortality rate ratios (Figure 3a). Among geriatric road users, states with several counties with a four-fold fatal mortality rate ratio were found in Oklahoma, Kansas, Texas, Utah, Nevada, California, and Oregon (Figure 3b). Among non-geriatric road users, states with higher than four-fold mortality rate ratio were found in almost all states in the West and South US regions as well as in North and South Dakota, Nebraska, and Kansas states (Figure 3c).

**Table 4:** Univariate association between county-level crash fatality and county multidimensional deprivation index, socioeconomic and crash characteristics

| Variables | Geriatric Road Users | Non-Geriatric Road Users | All Age Groups |
| --- | --- | --- | --- |
|  | Incidence Rate Ratio<br>(95% CrI) | Incidence Rate Ratio<br>(95% CrI) | Incidence Rate Ratio<br>(95% CrI) |
| MDI Rate (Median (Q1, Q3)) | 1.01 (1.01 – 1.02) | 1.01 (1.01 – 1.02) | 1.01 (1.01 – 1.01) |
| Deprivation Index (n (%)) |  |  |  |
| Very highly deprived | 1.34 (1.17 – 1.52) | 1.35 (1.20 – 1.50) | 1.33 (1.20 – 1.48) |
| Highly deprived | 1.21 (1.13 – 1.30) | 1.26 (1.18 – 1.34) | 1.25 (1.17 – 1.32) |
| Average to Low Deprived | Ref | Ref | Ref |
| <u>Sociodemographic Characteristics</u> |  |  |  |
| Male proportion | 1.07 (1.06 – 1.09) | 1.05 (1.04 – 1.06) | 1.05 (1.04 – 1.06) |
| Black proportion | 0.99 (0.99 – 0.99) | 0.99 (0.99 – 1.00) | 0.99 (0.99 – 1.00) |
| Hispanic proportion | 0.99 (0.99 – 0.99) | 0.99 (0.98 – 0.99) | 0.99 (0.99 – 0.99) |
| Poverty proportion | 1.03 (1.02 – 1.03) | 1.03 (1.02 – 1.03) | 1.02 (1.02 – 1.03) |
| Bachelor's degree proportion | 0.97 (0.97 – 0.97) | 0.96 (0.96 – 0.97) | 0.96 (0.96 – 0.97) |
| Excessive alcohol intake prop. | 0.00 (0.00 – 0.00) | 0.00 (0.00 – 0.00) | 0.00 (0.00 – 0.00) |
| ED utilization rate | 1.10 (1.07 – 1.14) | 1.10 (1.07 – 1.13) | 1.09 (1.06 – 1.11) |
| Rural + Small Towns | 1.81 (1.67 – 1.96) | 2.06 (1.94 – 2.20) | 1.97 (1.86 – 2.10) |
| Micropolitan Urban | 1.41 (1.32 – 1.50) | 1.52 (1.45 – 1.60) | 1.47 (1.40 – 1.54) |
| Metropolitan Urban | Ref | Ref | Ref |
| <u>Crash Event Characteristics</u> |  |  |  |
| Night fatal crash proportion | 0.99 (0.99 – 0.99) | 1.00 (0.99 – 1.00) | 0.99 (0.99 – 1.00) |
| Rush hour fatal event prop. | 1.00 (1.00 – 1.01) | 1.00 (1.00 – 1.00) | 1.00 (1.00 – 1.00) |
| Alcohol screening proportion | 0.99 (0.99 – 1.00) | 1.00 (1.00 – 1.00) | 1.00 (1.00 – 1.00) |
| Drug screening proportion | 0.99 (0.99 – 1.00) | 1.00 (1.00 – 1.00) | 1.00 (1.00 – 1.00) |
| <u>County Metrics (Coefficients)</u> |  |  |  |
| County: Random component | 11.93 (8.11 – 18.99) | 7.58 (6.67 – 9.70) | 9.15 (7.13 – 11.71) |
| County: Spatial component | 1.65 (1.27 – 2.07) | 1.40 (1.14 – 1.69) | 1.43 (1.18 – 1.71) |

MDI: Multidimensional Deprivation Index;

**Table 5:** Incidence rate ratios of crash fatalities involving older adults, teenagers, and all age groups across U.S. counties with varying levels of deprivation (N=3,108)

| Road Users | Variable | All Areas<br>(95% CrI) | Rural + Small Towns<br>(95% CrI) | Non-metropolitan<br>Urban (95% CrI) | Metropolitan<br>Urban (95% CrI) |
| --- | --- | --- | --- | --- | --- |
| Geriatric Road<br>Users (65+<br>years) | Deprivation Index |  |  |  |  |
|  | Very highly deprived | **1.23 (1.10 – 1.38)** | **1.46 (1.16 – 1.83)** | **1.44 (1.21 – 1.71)** | 1.17 (0.98 – 1.41) |
|  | Highly deprived | **1.15 (1.08 – 1.24)** | **1.28 (1.09 – 1.49)** | **1.26 (1.13 – 1.42)** | **1.19 (1.09 – 1.31)** |
|  | At or below the national<br>average | Ref | Ref | Ref | Ref |
| Non-Geriatric<br>Road Users | Deprivation Index |  |  |  |  |
|  | Very highly deprived | **1.20 (1.08 – 1.32)** | 1.20 (0.97 – 1.47) | **1.28 (1.10 – 1.48)** | **1.25 (1.05 – 1.48)** |
|  | Highly deprived | **1.19 (1.12 – 1.26)** | 1.13 (0.99 – 1.29) | **1.15 (1.05 – 1.27)** | **1.34 (1.23 – 1.47)** |
|  | At or below the national<br>average | Ref |  |  |  |
| All Age Groups | Deprivation Index |  |  |  |  |
|  | Very highly deprived | **1.20 (1.09 – 1.32)** | **1.22 (1.01 – 1.48)** | **1.29 (1.12 – 1.48)** | **1.23 (1.04 – 1.45)** |
|  | Highly deprived | **1.18 (1.12 – 1.25)** | **1.15 (1.02 – 1.30)** | **1.16 (1.06 – 1.27)** | **1.32 (1.21 – 1.44)** |
|  | At or below the national<br>average | Ref | Ref | Ref | Ref |

Model adjusted for male proportion, Hispanic proportion, the proportion of fatal crash events at night and during the rush hour period, the proportion of alcohol screening behavior at fatal crash events, and rurality/urbanicity. Rurality/urbanicity excluded from the rural, on-metropolitan urban, and metropolitan urban models since the variable was used for stratification.

**Figure 3:**
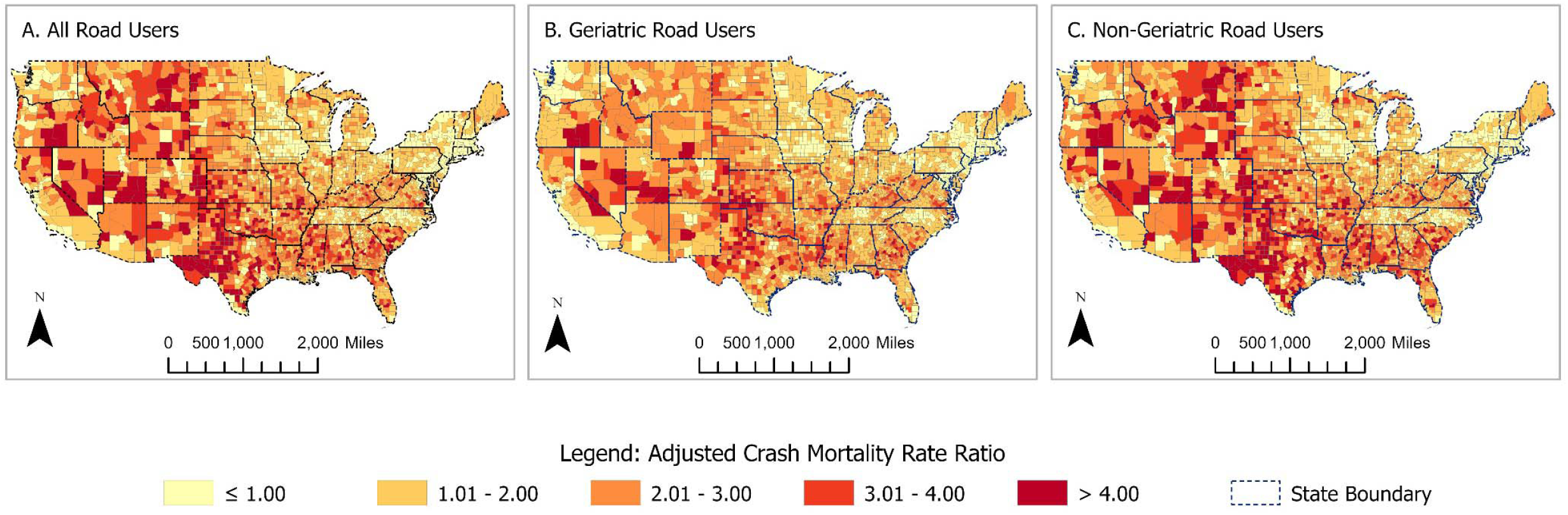
County-level distribution of adjusted crash mortality rates among (A) road users of all ages (B) geriatric road users, and (C) non-geriatric road users

## Discussion

Earlier studies have reported that social determinants of health are associated with cardiovascular,^28-30^ diabetes,^31, 32^ cancer,^33, 34^ and mental health-related morbidities and mortality.^35, 36^ Our study adds to the extant literature by reporting that social determinants of health are associated with fatal crash injuries among geriatric and non-geriatric road users. Our study validated that the multidimensional deprivation index is a composite measure that can be used to rank social determinants of health at the county level and we report that about a third of US counties had high to very high multidimensional deprivation indices. Rural-urban disparities exist in both the multidimensional deprivation index and fatal crash rates with rural areas having a larger proportion of counties with high and very high deprivation indices. Also, the geriatric and non-geriatric case fatality rates and SMR were highest in rural areas and lowest in metropolitan-urban areas. Counties with high and very high deprivation indices have significantly elevated fatal crash rate ratios and the ratios are not substantially different among geriatric and non-geriatric road users. Among geriatric road users, rural and micropolitan urban counties with high and very high deprivation indices have significantly elevated fatal crash rate ratios. However, among the non-geriatric road users, micropolitan-urban and metropolitan-urban counties with high and very high deprivation indices have significantly elevated fatal crash rate ratios. Lastly, we identified county hotspots of high deprivation indices as well as counties with more than four-fold increased fatality rate ratios in Texas, Oklahoma, Nevada, and Utah.

Although geriatric road users have higher case fatality rates and standardized mortality rates compared to non-geriatric road users, county-level measures of social determinants of health affect geriatric and non-geriatric road users equally. The elevated geriatric case fatality rate and standardized mortality rates may be a reflection of several factors such as the presence of co-morbidities,^37-40^, frailty,^41-46^, and under-appreciation and under-triage of the injury severity in the geriatric trauma population.^47-51^ However, the comparable effect of the multidimensional deprivation index on fatal crash injuries on geriatric and non-geriatric road users underscores the extent to which social determinants of health affect all road users, irrespective of age. This finding indicates that interventions targeting social determinants could be beneficial for all road users, regardless of age. Efforts to improve socioeconomic conditions, enhance educational opportunities, ensure equitable access to healthcare services, and create safe and supportive neighborhoods will, therefore, have a positive effect on the health and safety outcomes of both older adults and younger individuals. These findings emphasize the need for comprehensive approaches that address social determinants of health as a means of promoting road safety and well-being for all age groups.

We report that geriatric road users in rural counties have significantly higher fatality rate ratios while those geriatric road users in metropolitan-urban counties are at no significantly elevated crash fatality rate ratio. The significantly higher fatality rate ratios observed in rural counties suggest that there may be specific challenges unique to rural areas. Earlier studies have reported that residents in rural areas have longer emergency medical service (EMS) response times,^8, 10, 11^ limited access to level I or II trauma centers,^52-54^ and a higher proportion of deaths at the crash scene. In contrast, the lack of a significantly elevated crash fatality rate ratio among geriatric road users in metropolitan-urban counties suggests that factors such as better access to emergency medical services and more developed transportation networks, including helicopter EMS access, may contribute to improved injury outcomes for older adults in these areas. Addressing social determinants of health in rural areas may reduce fatal crash occurrences, especially among geriatric road users. Strategies to improve access to healthcare services, enhance transportation infrastructure, and create age-friendly environments can play a vital role in promoting road safety and protecting the well-being of geriatric road users. There is a need for collaborative efforts among policymakers, healthcare professionals, transportation agencies, and community stakeholders to develop and implement comprehensive interventions that address social determinants of health and improve road safety outcomes in rural areas.

Non-geriatric road users in metropolitan counties have significantly higher fatality rate ratios while those non-geriatric road users in rural counties are at no significantly elevated crash fatality rate ratio. This observed finding may suggest the disproportionate distribution of non-geriatric road users in metropolitan urban areas and the impact of risky driving behaviors associated with young and middle-aged drivers. Earlier studies have reported that non-geriatric road users are more likely to drive while under the influence of drugs and alcohol,^9, 55, 56^ and engage in phone-related distracted driving.^57-60^ These unique characteristics may account for the observed elevated fatal crash rate ratio in non-geriatric road users.

We identified fatality rate ratios in several counties located in Texas, Oklahoma, Nevada, and Utah. These states also have county hotspots of very high multidimensional deprivation index. Earlier studies have reported that these states have a high proportion of counties with prolonged EMS response time, county hotspots of rush hour-related fatal crash rates, and fatal crash rates due to non-use of seatbelt.^8, 14, 61, 62^ Identification of counties with high crash fatality rates and with high multidimensional deprivation index allows for focused interventions. Using spatial models and cluster analyses for fatal crash injury assessment can complement current efforts to address predictors of fatal crash injuries using the Motor Vehicle Prioritizing Interventions and Cost Calculator for States (MV PICCS).^63, 64^ Using the MV PICCS to identify which specific domain of social determinants of health will be the most cost-effective in reducing fatal crash injury at the state level and using spatial and spatiotemporal models to identify counties within the states that should receive the highest priority may be an effective and efficient way of addressing the social determinants of fatal crash injuries at the state and county-levels.

This study has its limitations. The ecological nature of the study makes it impossible to make causal inferences. We pooled our data from FARS, a data repository that relies on state-level reporting of crash events. Data entry and processing errors, as well as inconsistent reporting of crash and crash-related events across the US, cannot be eliminated. The MDI was computed from multiple data sources. Data errors intrinsic to each of the data are inherently transferred into the MDI computation and misclassification bias cannot be eliminated. Despite these limitations, this study is one of the few studies that assessed the association between social determinants of health and fatal crash rates among geriatric and non-geriatric road users. By identifying counties with high rates of fatal crash injury occurrences as those with a high level of deprivation, this study provides information that can guide targeted interventions to counties and states in greatest need. Additionally, our study provides information that can inform policy and resource allocation on improving the social determinants of health and preventing fatal crash occurrences in the presence of other competing community needs.

## Conclusion

Despite older adults being less frequent road users, geriatric road users have higher case fatality rates and standardized mortality rates. County-level deprivation measures of the SDoH are equally associated with geriatric and non-geriatric crash-related fatal rate ratios. Policies and interventions that improve county-level SDoH may reduce the county-level fatal rate ratios equally among geriatric and non-geriatric road users.

## Data Availability

All data produced in the present study are available upon reasonable request to the authors

## References

1. Braveman P, Egerter S, Williams DR. The social determinants of health: coming of age. Annu Rev Public Health. 2011;32:381–398.

2. Center for Disease Control and Prevention. Social Determinants of Health: Know What Affects Health. U.S. Department of Health and Human Services. Accessed 08/11/2020, https://www.cdc.gov/socialdeterminants/

3. Healthy People. Social determinants of health. Office of Disease Prevention and Health Promotion. Accessed 08/11/2020, https://www.healthypeople.gov/2020/topics-objectives/topic/social-determinants-of-health

4. Stajduhar KI, Mollison A, Giesbrecht M, et al. “Just too busy living in the moment and surviving”: barriers to accessing health care for structurally vulnerable populations at end-of-life. BMC Palliat Care. Jan 26 2019;18(1):11. doi:10.1186/s12904-019-0396-7

5. Bailey V. Rural Hospitals Facing Risk of Closure Due to Financial Stress. Revcycle Intelligence. Accessed 03/19/2023. https://revcycleintelligence.com/news/rural-hospitals-facing-risk-of-closure-due-to-financial-stress

6. Kaufman BG, Thomas SR, Randolph RK, et al. The Rising Rate of Rural Hospital Closures. The Journal of rural health. 2016;32(1):35–43. doi:10.1111/jrh.12128

7. Miller KEM, James HJ, Holmes GM, et al. The effect of rural hospital closures on emergency medical service response and transport times. 2020;55(2):288–300. doi:https://doi.org/10.1111/1475-6773.13254

8. Adeyemi OJ, Paul R, Arif A. An assessment of the rural-urban differences in the crash response time and county-level crash fatalities in the United States. The Journal of Rural Health. 2021/10/19 2021;doi:https://doi.org/10.1111/jrh.12627

9. Adeyemi OJ, Paul R, DiMaggio C, et al. Rush Hour-Related Road Crashes: Assessing the Social and Environmental Determinants of Fatal and Non-Fatal Road Crash Events. Ph.D. The University of North Carolina at Charlotte; 2021. https://www.proquest.com/dissertations-theses/rush-hour-related-road-crashes-assessing-social/docview/2572619606/se-2?accountid=14605

10. Adeyemi OJ, Paul R, DiMaggio C, et al. The association of crash response times and deaths at the crash scene: A cross-sectional analysis using the 2019 National Emergency Medical Service Information System. The Journal of Rural Health. 2022/04/22 2022;doi:https://doi.org/10.1111/jrh.12666

11. Byrne JP, Mann NC, Dai M, et al. Association Between Emergency Medical Service Response Time and Motor Vehicle Crash Mortality in the United States. JAMA surgery. 2019;154(4):286–293. doi:https://doi.org/10.1001/jamasurg.2018.5097

12. Adeyemi O. An Assessment of the Knowledge, Attitude, and Practice of Phone Use While Driving and Crash Outcomes Among Drivers in Oyo State, Nigeria. International Research and Review. 2022:25. https://files.eric.ed.gov/fulltext/EJ1334453.pdf

13. Adeyemi O. Alcohol- and Drug-Associated Injury Outcomes Among Older Adults Involved in Car Crashes. Innovation in aging. 2021;5(Suppl 1):127–127. doi:https://doi.org/10.1093/geroni/igab046.486

14. Adeyemi O, Paul R, Delmelle E, et al. Road environment characteristics and fatal crash injury during the rush and non-rush hour periods in the U.S: Model testing and cluster analysis. Spatial and Spatio-temporal Epidemiology. 2023/02/01/ 2023;44:100562. doi:https://doi.org/10.1016/j.sste.2022.100562

15. Saeednejad M, Sadeghian F, Fayaz M, et al. Association of Social Determinants of Health and Road Traffic Deaths: A Systematic Review. Bull Emerg Trauma. Oct 2020;8(4):211–217. doi:10.30476/beat.2020.86574

16. National Highway Traffic Safety Administration. Analytical User’s Manual 1975–2015. 2016;

17. National Highway Traffic Safety Administration. Fatalities and Fatality Rates by STATE, 1994 - 2018 - State : USA. Accessed 06/12/2020, https://www-fars.nhtsa.dot.gov/States/StatesFatalitiesFatalityRates.aspx

18. Glassman B. Producing County-level MDI Rates Using Public Use Data: 2010 to 2019. United States Census Bureau; 2022. Accessed 01/09/2023. https://www.census.gov/content/dam/Census/library/working-papers/2022/demo/sehsd-wp2022-19.pdf

19. Glassman B. Multidimensional Deprivation in the United States: 2017. ACS-40. United States Census Bureau; 2019. Accessed 06/23/2023. https://www.census.gov/library/publications/2019/acs/acs-40.html

20. Alkire S, Foster J. Counting and multidimensional poverty measurement. Journal of Public Economics. 2011/08/01/ 2011;95(7):476–487. doi:https://doi.org/10.1016/j.jpubeco.2010.11.006

21. United States Census Bureau. American Community Survey Data. Accessed 02/26/2020, 2020. https://www.census.gov/programs-surveys/acs/data.html

22. County Health Rankings and Roadmaps. Ranking data & documentation. Accessed July 14, 2020. https://www.countyhealthrankings.org/explore-health-rankings/rankings-data-documentation

23. University of Wisconsin Population Health Institute. 2020 County Health Rankings State Reports. COunty Health Ranking and Road Maps. 2020;(04/11/2020)

24. Agency for Healthcare Research and Quality. Healthcare Cost and Utilization Project. Accessed 06/24/2023, https://hcup-us.ahrq.gov/databases.jsp

25. Adeyemi OJ, Arif AA, Paul R. Exploring the relationship of rush hour period and fatal and non-fatal crash injuries in the U.S.: A systematic review and meta-analysis. Accid Anal Prev. Oct 28 2021;163:106462. doi:10.1016/j.aap.2021.106462

26. Economic Research Services. Rural-Urban Commuting Area Codes. 2019;(04/11/2020)

27. Ke W, Cheng HP, Yan D, et al. The application of cluster analysis and inverse distance-weighted interpolation to appraising the water quality of Three Forks Lake. Procedia Environmental Sciences. 2011;10:2511–2517.

28. Powell-Wiley TM, Baumer Y, Baah FO, et al. Social Determinants of Cardiovascular Disease. Circ Res. Mar 4 2022;130(5):782–799. doi:10.1161/circresaha.121.319811

29. Kimenai DM, Pirondini L, Gregson J, et al. Socioeconomic Deprivation: An Important, Largely Unrecognized Risk Factor in Primary Prevention of Cardiovascular Disease. Circulation. 2022/07/19 2022;146(3):240–248. doi:10.1161/CIRCULATIONAHA.122.060042

30. Flynn A, Vaughan A, Casper M, et al. Differences in Heart Disease Death Rates and Trends by County-Level Deprivation, 2010-2019. United States Census BUreau; 2021. Accessed 01/09/2023. https://www.census.gov/content/dam/Census/library/working-papers/2021/demo/sehsd-wp2021-19.pdf

31. Butler AM. Social Determinants of Health and Racial/Ethnic Disparities in Type 2 Diabetes in Youth. Curr Diab Rep. Aug 2017;17(8):60. doi:10.1007/s11892-017-0885-0

32. Walker RJ, Strom Williams J, Egede LE. Influence of Race, Ethnicity and Social Determinants of Health on Diabetes Outcomes. Am J Med Sci. Apr 2016;351(4):366–73. doi:10.1016/j.amjms.2016.01.00833.

33. Coughlin SS. Social determinants of breast cancer risk, stage, and survival. Breast Cancer Res Treat. Oct 2019;177(3):537–548. doi:10.1007/s10549-019-05340-7

34. Coughlin SS. Social determinants of colorectal cancer risk, stage, and survival: a systematic review. Int J Colorectal Dis. Jun 2020;35(6):985–995. doi:10.1007/s00384-020-03585-z

35. Alegría M, NeMoyer A, Falgàs Bagué I, et al. Social Determinants of Mental Health: Where We Are and Where We Need to Go. Current psychiatry reports. 2018;20(11):95–95. doi:10.1007/s11920-018-0969-9

36. Allen J, Balfour R, Bell R, et al. Social determinants of mental health. Int Rev Psychiatry. Aug 2014;26(4):392–407. doi:10.3109/09540261.2014.928270

37. Salive ME. Multimorbidity in older adults. Epidemiol Rev. 2013;35:75–83. doi:10.1093/epirev/mxs009

38. Divo MJ, Martinez CH, Mannino DM. Ageing and the epidemiology of multimorbidity. Eur Respir J. Oct 2014;44(4):1055–68. doi:10.1183/09031936.00059814

39. Shigemura N, Toyoda Y. Elderly patients with multiple comorbidities: insights from the bedside to the bench and programmatic directions for this new challenge in lung transplantation. Transpl Int. Apr 2020;33(4):347–355. doi:10.1111/tri.13533

40. Davis JW, Chung R, Juarez DT. Prevalence of comorbid conditions with aging among patients with diabetes and cardiovascular disease. Hawaii Med J. Oct 2011;70(10):209–13.

41. Adeyemi O, Grudzen C, DiMaggio C, et al. Pre-Injury Frailty and Clinical Care Trajectory of Geriatric Trauma Patients: A Retrospective Cohort Analysis of A Large Level I US Trauma Center. medRxiv. 2023;doi:https://doi.org/10.1101/2023.06.19.23291575

42. Bandeen-Roche K, Seplaki CL, Huang J, et al. Frailty in Older Adults: A Nationally Representative Profile in the United States. The Journals of Gerontology: Series A. 2015;70(11):1427–1434. doi:10.1093/gerona/glv133

43. Chen X, Mao G, Leng SX. Frailty syndrome: an overview. Clin Interv Aging. 2014;9:433–41. doi:10.2147/cia.S45300

44. Curtis E, Romanowski K, Sen S, et al. Frailty score on admission predicts mortality and discharge disposition in elderly trauma patients over the age of 65 y. J Surg Res. Oct 2018;230:13–19. doi:10.1016/j.jss.2018.04.017

45. Fehlmann CA, Nickel CH, Cino E, et al. Frailty assessment in emergency medicine using the Clinical Frailty Scale: a scoping review. Intern Emerg Med. Nov 2022;17(8):2407–2418. doi:10.1007/s11739-022-03042-5

46. Lin HS, Watts JN, Peel NM, et al. Frailty and post-operative outcomes in older surgical patients: a systematic review. BMC Geriatr. Aug 31 2016;16(1):157. doi:https://doi.org/10.1186/s12877-016-0329-8

47. Alshibani A, Alharbi M, Conroy S. Under-triage of older trauma patients in prehospital care: a systematic review. Eur Geriatr Med. 2021;12(5):903–919. doi:https://doi.org/10.1007/s41999-021-00512-5

48. Anantha RV, Painter MD, Diaz-Garelli F, et al. Undertriage Despite Use of Geriatric-Specific Trauma Team Activation Guidelines: Who Are We Missing? Am Surg. 2021/03/01 2020;87(3):419–426. doi:https://doi.org/10.1177/0003134820951450

49. Chang DC, Bass RR, Cornwell EE, et al. Undertriage of elderly trauma patients to state-designated trauma centers. Arch Surg. Aug 2008;143(8):776–81; discussion 782. doi:https://doi.org/10.1001/archsurg.143.8.776

50. Chu I, Vaca F, Stratton S, et al. Geriatric trauma care: challenges facing emergency medical services. California Chapter of the American Academy of Emergency Medicine. Updated 06/12/2022. Accessed 04/13/2022, https://www.ncbi.nlm.nih.gov/pmc/articles/PMC2860422/pdf/cjem8_2p0051.pdf

51. Horst MA, Morgan ME, Vernon TM, et al. The geriatric trauma patient: A neglected individual in a mature trauma system. J Trauma Acute Care Surg. Jul 2020;89(1):192–198. doi:10.1097/ta.0000000000002646

52. Dave U, Gosine B, Palaniappan A. An Overview of Trauma Center Levels and Disparities in Rural Trauma Care. Reconstructive Review. 2020;10doi:https://doi.org/10.15438/rr.10.1.234

53. Hsia R, Shen YC. Possible geographical barriers to trauma center access for vulnerable patients in the United States: an analysis of urban and rural communities. Arch Surg. Jan 2011;146(1):46–52. doi:10.1001/archsurg.2010.299

54. Newgard CD, Fu R, Bulger E, et al. Evaluation of Rural vs Urban Trauma Patients Served by 9-1-1 Emergency Medical Services. JAMA Surgery. 2017;152(1):11–18. doi:10.1001/jamasurg.2016.3329

55. Li G, Brady JE, Chen Q. Drug use and fatal motor vehicle crashes: a case-control study. Accid Anal Prev. Nov 2013;60:205–10. doi:10.1016/j.aap.2013.09.001

56. Martin J-L, Gadegbeku B, Wu D, et al. Cannabis, alcohol and fatal road accidents. PLoS One. 2017;12(11):e0187320. doi:10.1371/journal.pone.0187320

57. Bergmark RW, Gliklich E, Guo R, et al. Texting while driving: the development and validation of the distracted driving survey and risk score among young adults. Inj Epidemiol. Dec 2016;3(1):7. doi:10.1186/s40621-016-0073-8

58. Boccardi V, Paolisso G. Distracted driving and crash risk. The New England Journal of Medicine. 2014;370(16):1565–1565. doi:10.1056/NEJMc1401241

59. Coben JH, Zhu M. Keeping an eye on distracted driving. JAMA: Journal of the American Medical Association. 2013;309(9):877–878. doi:10.1001/jama.2013.491

60. Foss RD, Goodwin AH. Distracted driver behaviors and distracting conditions among adolescent drivers: Findings from a naturalistic driving study. J Adolesc Health. 2014;54(5, Suppl):S50–S60. doi:10.1016/j.jadohealth.2014.01.005

61. Adeyemi O, Paul R, Arif A. Spatial Cluster Analysis of Fatal Road Accidents From Non-Use of Seat Belts Among Older Drivers. Innovation in Aging. 2020;4(Supplement_1):113–114. doi:https://doi.org/10.1093/geroni/igaa057.374

62. Adeyemi O, Rajib P, Arif A. Rush hour accidents: Assessing the relationship between road environmental determinants and county-level fatal road accidents patterns in the United States. Accessed 10/11/2021, https://apha.confex.com/apha/2020/meetingapp.cgi/Paper/483798

63. Center for Disease Control and Prevention. Motor Vehicle Prioritizing Interventions and Cost Calculator for States (MV PICCS). National Center for Injury Prevention and Control. Accessed 12/8/2020, https://www.cdc.gov/transportationsafety/calculator/index.html

64. Ecola L, Ringel JS, Connor K, et al. Costs and Effectiveness of Interventions to Reduce Motor Vehicle-Related Injuries and Deaths: Supplement to Tool Documentation. Rand health quarterly. 2018;8(2):9–9.

